# Phenome-Wide Association Study of Pre-Cancer Diagnosis Electronic Health Records Identifies Risk and Inverse Associations in the All of Us Research Program

**DOI:** 10.64898/2026.05.26.26353823

**Authors:** C. Christian D. Rich, Ethan J. Bang, Alyssa B. Bair, Britton E. Richardson, Jocelyn L. Millington, Blaine A. Bates, Mary F. Davis, Matthew H. Bailey

## Abstract

**Background:** The *All of Us* Research Program represents a rich resource for cancer epidemiology research, with over 400,000 participants with whole genome sequences linked to electronic health records (EHR). Large cancer datasets often focus exclusively on cases without controls and neglect pre-diagnosis healthcare occurrences. Here, we perform a phenome-wide association study (PheWAS) of EHR data at least 1 year pre-diagnosis between cancer cases and matched controls, revealing co-occurring and mutually exclusive phenotypes.

**Methods:** We identified 55,000+ cancer cases across 21 cancer types in *All of Us* version 8. To eliminate age-related confounding, we implemented a two-stage matching and censoring strategy: loose matching on demographics to establish index dates and cohort comparability, followed by right-censoring of EHR data (excluding 1 year pre-diagnosis/index), then 1:2 matching to address residual demographic imbalance. We tested associations between 23,193 cancer cases, 46,386 matched controls and approximately 1,600 clinical phenotypes using logistic regression adjusted for sex at birth, self-reported race, age at diagnosis/index date, and two censored EHR metrics: observation window and unique condition count, with Bonferroni correction for multiple testing.

**Results:** Our analysis identified 232 significantly associated phenotypes, confirming established cancer risk factors including elevated prostate specific antigen (OR = 2.92, 95% CI: 2.65-3.23; p-value=1.8×10-101) and multinodular goiter (OR = 1.73, 95% CI: 1.56-1.91; p-value = 6.7×10-27). Further investigation into the relationship between several phenotypes with seeming inverse effects is warranted.

**Conclusions:** This PheWAS of EHR data at least 1 year pre-diagnosis leveraged the diversity of *All of Us* to examine how clinical phenotypes prior to cancer diagnosis vary across cancer types and racial groups. Our findings validate *All of Us* as a robust platform for cancer epidemiology research, confirming established risk factors at scale across diverse populations. This work provides methodological insights for EHR-based susceptibility analyses and demonstrates the value of agnostic phenome-wide approaches for generating hypotheses in precision medicine.

**Graphical Abstract:** 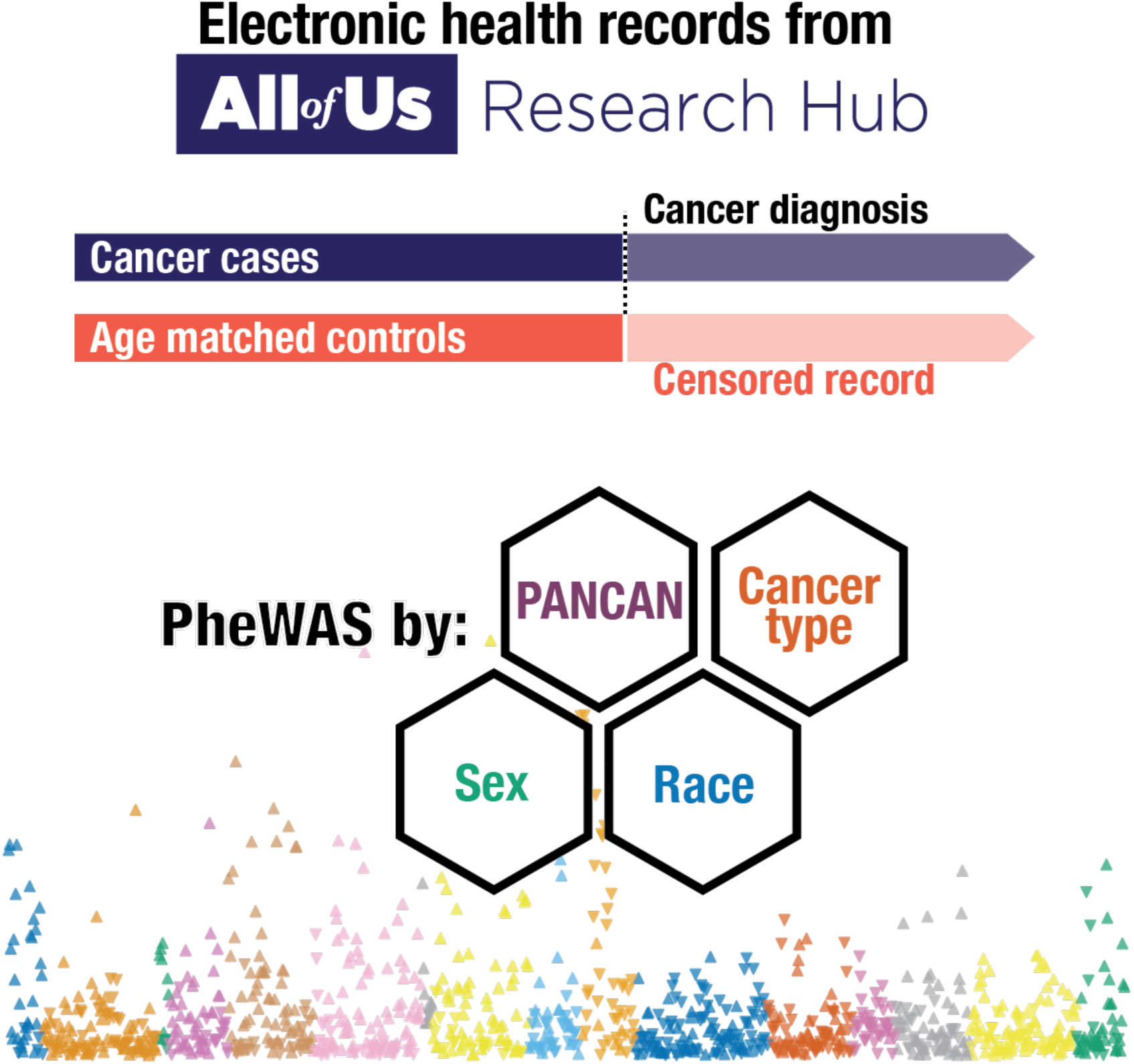

Using EHR data from over 69,000 participants, this study leveraged the diversity of *All of Us* to examine how clinical phenotypes prior to cancer diagnosis vary across cancer types and racial groups.

## Introduction

Modern precision medicine aims to tailor healthcare interventions based on individual characteristics, requiring comprehensive understanding of how genetic, environmental, and clinical factors interact to influence disease risk and outcomes.^1,2^ To support this vision, large-scale biobanks have emerged that link longitudinal electronic health records (EHR) and enable researchers to explore disease relationships agnostically at scale.^3,4^ Phenome-wide association studies (PheWAS) leverage this data to test for associations between genetic variants and clinical phenotypes (phenotype ∼ genotype), replicating known gene-disease relationships and identifying unexpected associations.^5,6^

Cancer remains a leading cause of death in the United States, with over 2 million new diagnoses and more than 600,000 deaths expected in 2026.^7^ Traditional cancer research has focused primarily on identifying risk factors that increase cancer likelihood, leading to well-established prevention strategies targeting modifiable behaviors and environmental exposures.^8–10^ Yet gaps remain in our understanding of the cancer phenome, particularly in diverse populations that have been historically underrepresented in large-scale genomic studies.^11–13^

The *All of Us* Research Program (*All of Us)* represents an unprecedented initiative to advance precision medicine through comprehensive data collection from diverse populations across the United States.^3^ As of the most recent data release (version 8, February 5, 2025), *All of Us* includes more than 600,000 participants with longitudinal EHR and demographic data.^14^ Importantly, the cohort’s size and population diversity provides sufficient cancer cases across multiple cancer types alongside controls. Large-scale cancer genomics initiatives such as The Cancer Genome Atlas have advanced understanding of tumor biology through case-only designs.^15^ While these efforts have been transformative for understanding cancer heterogeneity, they do not address the pre-cancer phenome that may signal cancer risk.

Cancer offers a compelling application for the PheWAS framework given its complex etiology involving genetic, environmental, and stochastic factors.^16^ We use the PheWAS approach to examine cancer as the independent variable of interest, modeling each clinical phenotype as a function of cancer diagnosis plus covariates (phenotype ∼ cancer + covariates). This disease-disease framework complements targeted investigation by scanning for pre-existing conditions that associate with subsequent cancer diagnosis, generating hypotheses for understanding cancer susceptibility.^17,18^

Here, we present a PheWAS of EHR data at least 1 year pre-diagnosis examining associations between malignant cancer diagnosis (hereafter “cancer”) and over 1,600 clinical phenotypes in *All of Us*. A key challenge for case-control analyses is differential observation periods, which has led prior studies to focus on prognostic outcomes for cases rather than pre-diagnosis comparisons between cases and controls.^19^ Recent disease association studies have attempted to address this by censoring case data and recalculating “the ages of the subjects… to reflect the censoring”.^20,21^ Building on this approach, we implemented a two-stage matching and censoring strategy. First, we performed loose matching on birth year, sex, and race to assign each control an index date corresponding to their matched case’s diagnosis date. This initial matching adjusts for cohort effects by ensuring cases and controls are drawn from similar birth cohorts. We then right censored all EHR data, excluding records occurring within 1 year before the index date. Following censoring, we performed a second round of 1:2 matching on birth year, age at index date, sex, and race to ensure cases and controls are at risk for outcomes during the same age ranges. We conducted both pan-cancer and cancer type specific analyses for 23,193 cancer cases across 21 cancer types matched to 46,386 controls. Our findings validate *All of Us* as a robust platform for cancer epidemiology research, confirming established risk factors at scale across diverse populations. This work provides methodological insights for EHR-based susceptibility analyses and demonstrates the value of agnostic phenome-wide approaches for generating hypotheses in precision medicine.

## Results

### Study Population and Cohort Characteristics

From the *All of Us* version 8 Controlled Tier Dataset containing over 390,000 participants with EHR data, we identified 55,800 individuals with at least one cancer-diagnosis Systematized Nomenclature of Medicine—Clinical Terms (SNOMED CT) code. After applying inclusion criteria requiring documented sex at birth and race information, restriction to unrelated individuals, cancer cases with compatible EHR data at least 1 year pre-diagnosis, and controls without any malignant-related diagnoses, 29,806 cancer cases and 203,911 controls remained eligible for matched case-control analysis (*n* = 233,717).

Following a two-stage matched case-control design to align temporal EHR censoring, the final analytic cohort consisted of 69,579 participants, with a 1:2 ratio of cases (*n* = 23,193) to controls (*n* = 46,386) (Fig. 1). The cohort was 63.0% female (n = 43,821) and 37.0% male (n = 25,758). Self-reported race distribution included 70.1% Non-Hispanic White (n = 48,804), 15.5% Non-Hispanic Black (n = 10,764), 11.3% Hispanic (n = 7,872), 1.8% Asian (n = 1,236), 0.8% American Indian or Alaska Native (n = 528), 0.4% Middle Eastern or North African (n = 291), and 0.1% Native Hawaiian or Other Pacific Islander (n = 84) participants. Mean age at censoring was 58.9 years (SD = 12.5) for both cases and controls, with all demographic and EHR-related measurement distributions presented in Table 1.

**Fig 1.**
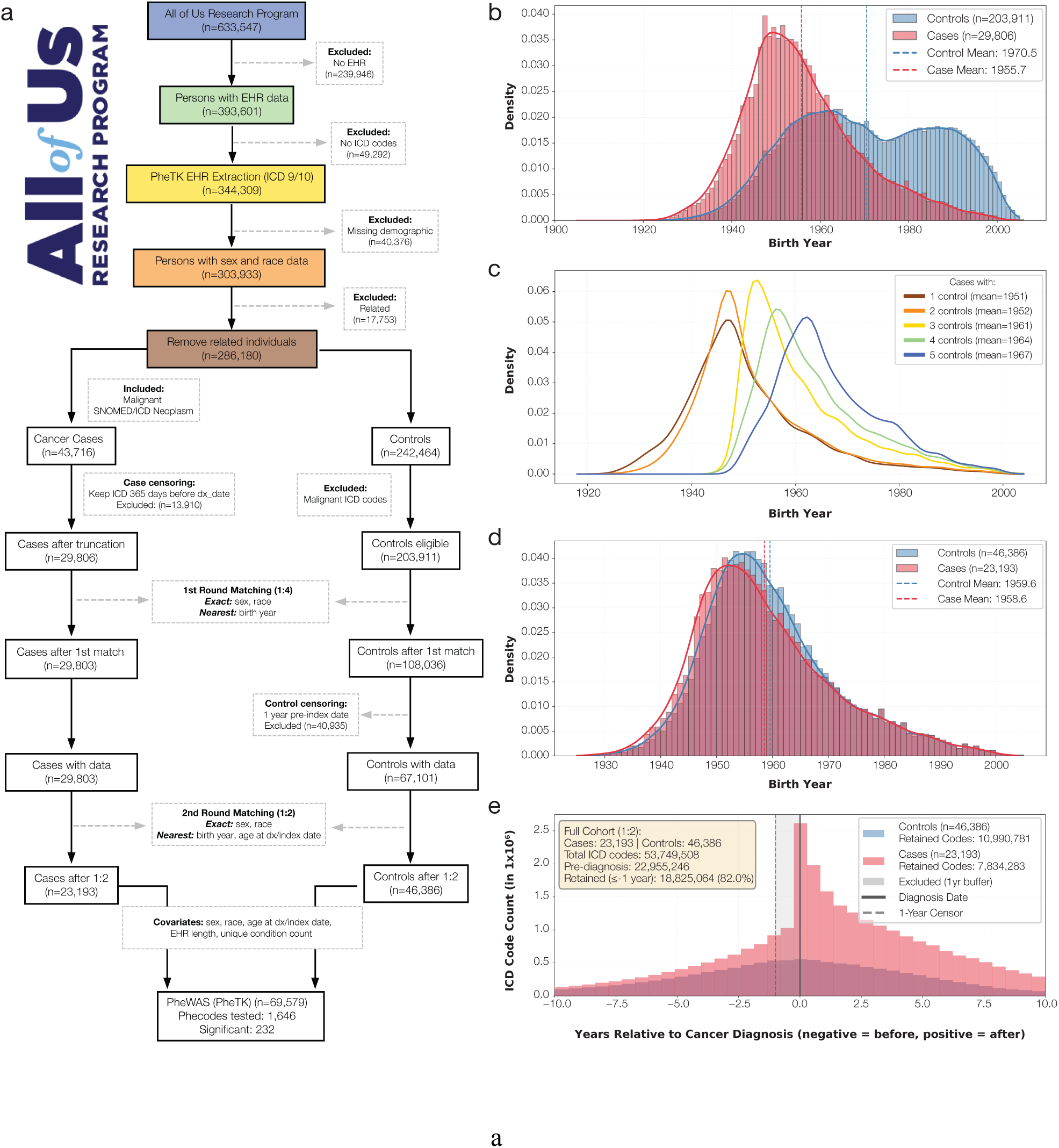
Study cohort selection and demographic balancing through sequential matching and temporal censoring. **a**, CONSORT flow diagram showing selection from *All of Us* participants (n=633,537) to the final PheWAS analytic cohort (n=69,579) after applying eligibility criteria, cancer case identification, and two-stage matching. **b**, Birth year density distributions before matching, showing age imbalance between cancer cases (mean: 1955.7 years) and controls (mean: 1970.5 years). **c**, Birth year distribution after first-round variable-ratio matching, stratified by number of controls per case, showing residual imbalance. **d**, Birth year density distributions after second-round 1:2 matching, demonstrating balance with mean birth year difference of 1.0 year (cases: 1958.6 vs. controls: 1959.6). **e**, ICD code timing distribution relative to cancer diagnosis/index date, normalized by sample size (2:1 control to case ratio). Prominent spike at year 0 for cases (pink) reflects diagnostic workup codes. Dashed vertical black line at −1 year indicates censoring threshold; gray shaded region shows excluded codes. Of 53,749,508 total ICD codes, 18,825,064 codes >1 year pre-diagnosis were retained (cases: 128.5 codes/person; controls: 112.4 codes/person).

**Table 1:** Demographic and clinical characteristics of the final matched cohort. Demographics include age at diagnosis/index date, birth year, sex at birth, and self-reported race. Clinical characteristics include censored EHR observation window, unique condition count, and total code occurrence count. AIAN = American Indian/Alaska Native; MENA = Middle Eastern/North African; NHPI = Native Hawaiian/Pacific Islander.

| Characteristic | Overall (N=69,579) | Cases (N=23,193) | Controls (N=46,386) |
| --- | --- | --- | --- |
| DEMOGRAPHICS |  |  |  |
| Age at Diagnosis/Index Date, years |  |  |  |
| Mean (SD) | 58.9 (12.5) | 58.9 (12.7) | 58.9 (12.3) |
| Median [Range] | 60.7 [7.5-98.7] | 60.8 [7.5-95.8] | 60.6 [9.8-98.7] |
| Birth Year |  |  |  |
| Mean (SD) | 1959.2 (12.0) | 1958.6 (12.3) | 1959.6 (11.8) |
| Median [Range] | 1957 [1925-2004] | 1956 [1927-2004] | 1958 [1925-2004] |
| Sex at birth, N (%) |  |  |  |
| Female | 43,821 (63.0%) | 14,607 (63.0%) | 29,214 (63.0%) |
| Male | 25,758 (37.0%) | 8,586 (37.0%) | 17,172 (37.0%) |
| Self-Reported Race,<br>N (%) |  |  |  |
| Non-Hispanic White | 48,804 (70.1%) | 16,268 (70.1%) | 32,536 (70.1%) |
| Non-Hispanic Black | 10,764 (15.5%) | 3,588 (15.5%) | 7,176 (15.5%) |
| Hispanic | 7,872 (11.3%) | 2,624 (11.3%) | 5,248 (11.3%) |
| Asian | 1,236 (1.8%) | 412 (1.8%) | 824 (1.8%) |
| AIAN | 528 (0.8%) | 176 (0.8%) | 352 (0.8%) |
| MENA | 291 (0.4%) | 97 (0.4%) | 194 (0.4%) |
| NHPI | 84 (0.1%) | 28 (0.1%) | 56 (0.1%) |
| **EHR CHARACTERISTICS** |  |  |  |
| EHR Observation<br>Window, years |  |  |  |
| Mean (SD) | 7.0 (7.4) | 7.4 (7.3) | 6.8 (7.4) |
| Median [Range] | 4.9 [0.0-56.4] | 5.3 [0.0-56.4] | 4.6 [0.0-42.4] |
| Total Unique Conditions |  |  |  |
| Mean (SD) | 39.6 (50.3) | 47.9 (57.2) | 35.5 (45.9) |
| Median [Range] | 21 [1-663] | 28 [1-616] | 19 [1-663] |
| Total ICD Code<br>Occurrences |  |  |  |
| Mean (SD) | 128.5 (262.7) | 160.6 (312.1) | 112.4 (232.5) |
| Median [Range] | 42 [1-9658] | 57 [1-9658] | 36 [1-9543] |

### Pan-cancer PheWAS Analysis

We conducted PheWAS using EHR data at least 1 year pre-diagnosis to identify clinical phenotypes associated with cancer. In the primary pan-cancer analysis, the final cohort contributed 3,258 unique phecodeX phenotypes across 3,282,591 total phecode events. After excluding those with fewer than 20 cases or controls, 2,317 phecodes were tested for association with cancer diagnosis using multivariable logistic regression. Of 1,646 converged tests, 232 phecodes exceeded the Bonferroni-corrected significance threshold (*p-*value < 3.04×10⁻⁵) (Fig. 2). Significant associations clustered within several clinical categories: gastrointestinal (*n* = 39), genitourinary (*n* = 35), neoplasms (*n* = 28), endocrine/metab (*n* = 26), blood/immune (*n* = 25), dermatological (*n* = 23). Complete results from all PheWAS analyses, including effect estimates, confidence intervals, p-values, and case counts for each phecode-cancer association, are provided in Supplementary Data 1.

**Fig 2.**
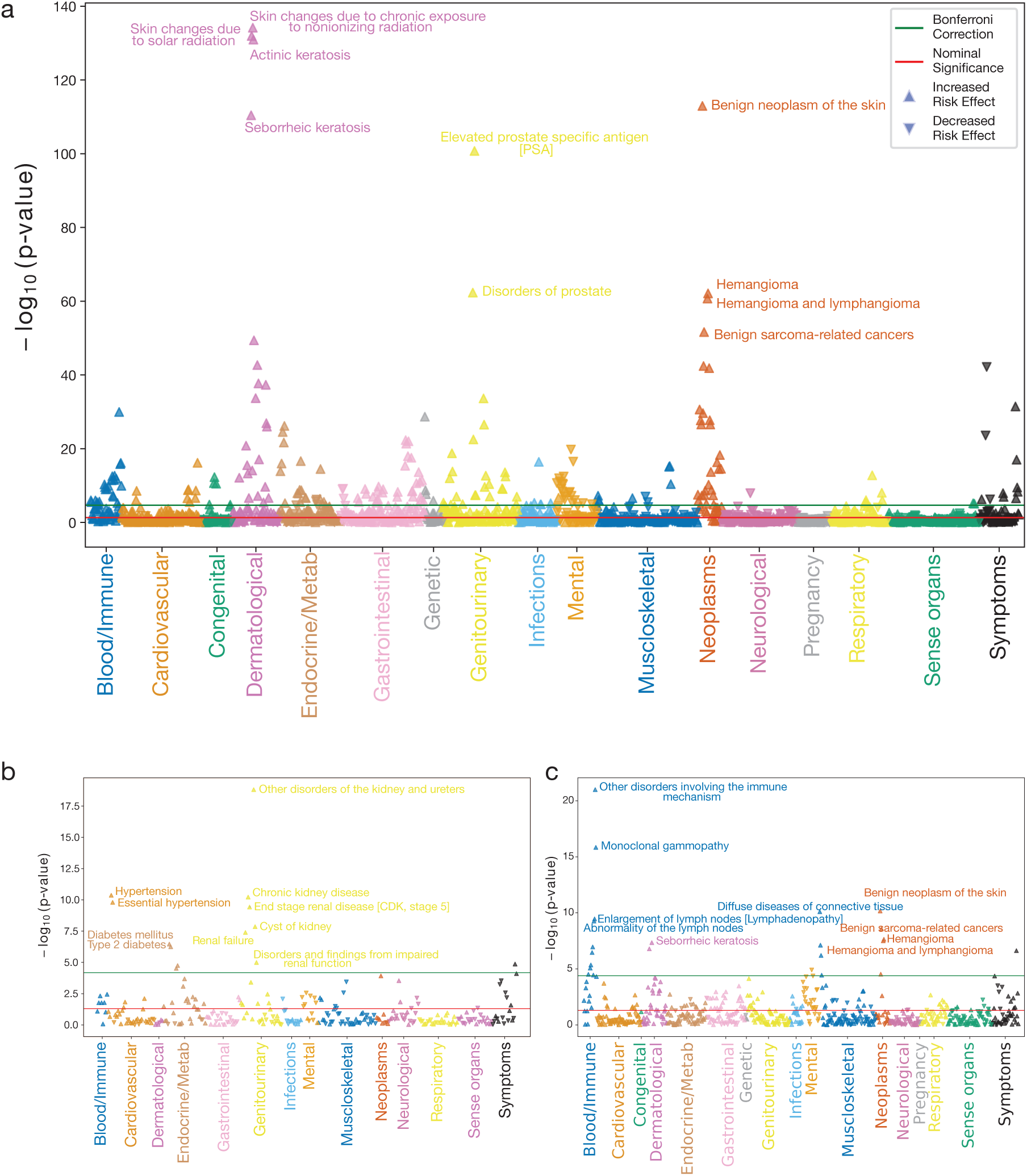
Phenome-wide associations between clinical phenotypes and cancer diagnosis. Manhattan plots showing −log₁₀(p-value) for associations between phecodeX phenotypes (x-axis, color-coded by disease category) and cancer diagnosis. Red horizontal line indicates p = 0.05; green line indicates Bonferroni-corrected significance threshold. Upward triangles represent increased risk associations; downward triangles represent inverse associations (legend applies to all panels). **a**, Pan-cancer analysis (all cancer types combined; cases: *n* = 23,193, controls: *n =* 46,386). **b**, Kidney cancer-specific analysis (cases: *n* = 804, controls: *n* = 1608). **c**, Blood cancer-specific analysis (leukemia, non-Hodgkin’s lymphoma, and myeloma combined; cases: *n* = 2,136, controls: *n* = 4,272). Each analysis applied independent Bonferroni correction based on phenotypes tested within that stratum.

Dermatological conditions exhibited the strongest positive associations. Of these, actinic keratosis demonstrated the highest odds ratio (OR = 2.62, 95% CI: 2.42-2.83; *p-*value = 1.2×10^-131^), while skin changes due to chronic exposure to nonionizing radiation (OR = 2.42, 95% CI: 2.25-2.59; *p-*value = 6.8×10^-135^) and skin changes due to solar radiation (OR = 2.31, 95% CI: 2.16-2.47; *p-*value = 1.2×10^-132^) showed strong associations.

In females, breast conditions such as benign mammary dysplasias (OR = 1.60, 95% CI: 1.48-1.73; *p-*value = 2.4×10^-34^) and lump or mass in breast or nonspecific abnormal breast exam (OR = 1.45, 95% CI: 1.38-1.53; *p-*value = 1.4×10^-42^) were notable associations. Male-specific associations included elevated prostate-specific antigen (OR = 2.92, 95% CI: 2.65-3.23; *p-*value = 1.8×10^-101^) and benign prostatic hyperplasia (OR = 1.42, 95% CI: 1.33-1.53; *p-*value = 3.2×10^-23^). Thyroid disorders, including multinodular goiter (OR = 1.73, 95% CI: 1.56-1.91; *p-*value = 6.7×10^-27^), also showed significant associations.

### Cancer Type Specific Associations

Cancer type specific PheWAS revealed associations unique to individual cancer types alongside shared risk factors observed in pan-cancer analysis (Fig. 2). In kidney cancer, the associations included chronic kidney disease (OR = 2.75, 95% CI: 2.03-3.72; *p-*value = 6.0×10^-11^) and other disorders of the kidney and ureters (OR = 5.54, 95% CI: 3.82-8.02; *p-*value = 1.5×10^-19^), as well as type 2 diabetes (OR = 1.71, 95% CI: 1.39-2.11; *p-*value = 5.6×10^-7^) and transplanted organ (OR = 4.84, 95% CI: 2.38-9.86; *p-*value = 1.4×10^-5^). For blood cancers (leukemia, non-Hodgkin’s lymphoma, and myeloma), immune-related phenotypes showed the largest effect sizes, such as other disorders involving the immune mechanism (OR = 5.19, 95% CI: 3.71-7.28; *p-*value = 1.0×10^-21^), abnormality of the lymph nodes (OR = 2.08, 95% CI: 1.65-2.62; *p-*value = 5.5×10^-10^) and monoclonal gammopathy (OR = 11.19, 95% CI: 6.31-19.85; *p-*value = 1.4×10^-16^).

### Race Stratified Analyses

Race stratified analyses revealed both shared risk factors and population-specific disease associations (Fig. 3). Among Non-Hispanic Black participants, the strongest positive associations included disorders of the prostate (OR = 2.28, 95% CI: 1.92-2.70; *p-*value = 5.6×10^-21^), elevated prostate-specific antigen (OR = 4.20, 95% CI: 3.18-5.55; *p-*value = 4.1×10^-24^), and benign mammary dysplasias (OR = 1.86, 95% CI: 1.50-2.30; *p-*value = 1.3×10^-8^). Notably, multiple anemia-related phenotypes showed significant associations, including deficiency anemias (OR = 1.33, 95% CI: 1.17-1.53; *p-*value = 2.7×10^-5^) and anemia (OR = 1.31, 95% CI: 1.17-1.47; *p-*value = 2.7×10^-6^).

**Fig 3.**
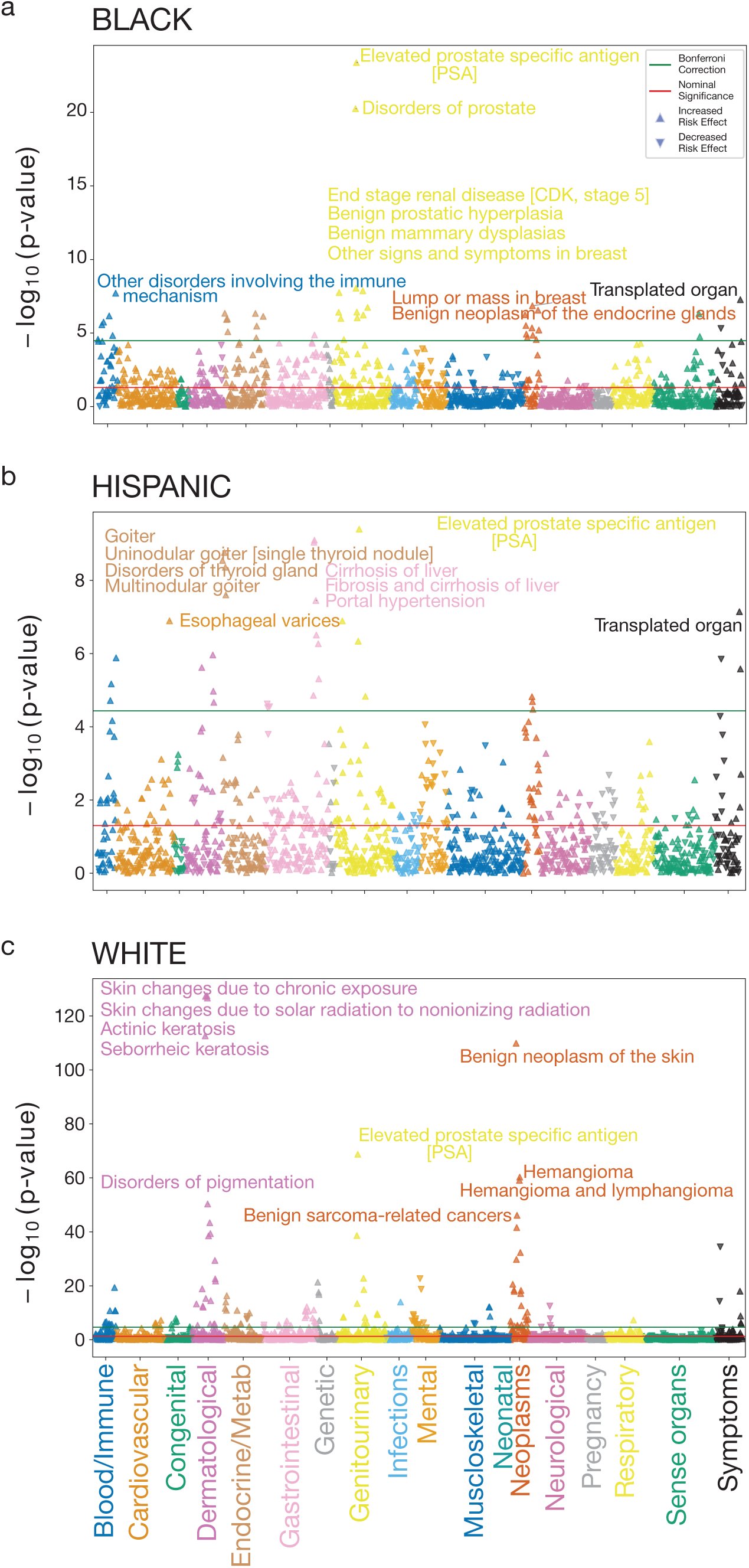
Race stratified phenome-wide associations between clinical phenotypes and cancer diagnosis. Manhattan plots showing −log₁₀(p-value) for associations between phecodeX phenotypes (x-axis, color-coded by disease category) and cancer diagnosis. Red horizontal line indicates p = 0.05; green line indicates Bonferroni-corrected significance threshold. Upward triangles represent increased risk associations; downward triangles represent inverse associations (legend applies to all panels). **a**, Non-Hispanic Black participants (cases: *n* = 3,588, controls: *n* = 7,176). **b**, Hispanic participants (cases: *n* = 2,624, *n* = 5,248). **c**, Non-Hispanic White participants (cases: *n* = 16,268, controls: *n* = 32,536). Each analysis applied independent Bonferroni correction based on phenotypes tested within that stratum.

For Hispanic participants, liver-related phenotypes featured prominently, including chronic nonalcoholic liver disease (OR = 1.66, 95% CI: 1.37-2.02; *p-*value = 3.1×10^-7^), portal hypertension (OR = 4.07, 95% CI: 2.47-6.71; *p-*value = 3.5×10^-8^), and cirrhosis of liver (OR = 2.59, 95% CI: 1.91-3.50; *p-*value = 7.9×10^-10^). Thyroid disorders including multinodular goiter (OR = 2.60, 95% CI: 1.86-3.64; *p-*value = 2.5×10^-8^) and uninodular goiter (OR = 2.30, 95% CI: 1.74-3.03; *p-*value = 4.4×10^-9^) also showed significant associations.

Among Non-Hispanic White participants, skin changes due to chronic exposure to nonionizing radiation (OR = 2.42, 95% CI: 2.25-2.60; *p-*value = 3.6×10^-128^) and seborrheic keratosis (OR = 2.59, 95% CI: 2.38-2.81; *p-*value = 2.4×10^-113^) demonstrated robust effect sizes. Benign breast conditions in females and prostate-related conditions in males remained strongly associated, consistent with pan-cancer and other race stratified results.

### Phenotype-Specific Patterns Across Cancer Types

Beyond cancer type and race-stratified patterns, individual phenotypes demonstrated heterogeneous associations across the cancer spectrum. For instance, dermatological conditions showed varying effects across different cancer types, while transplanted organ exhibited a strong association with kidney cancer (Fig. 4). Inverse associations for conditions including chronic pain (OR = 0.70, 95% CI: 0.67-0.74; *p-*value = 6.7×10^-43^) and psychiatric-related diagnoses such as mood disorders (OR = 0.82, 95% CI: 0.78-0.85; *p-*value = 1.6×10^-20^) were consistent across most cancer types, with strong effects observed for skin and breast cancers (Fig. 5).

**Fig 4.**
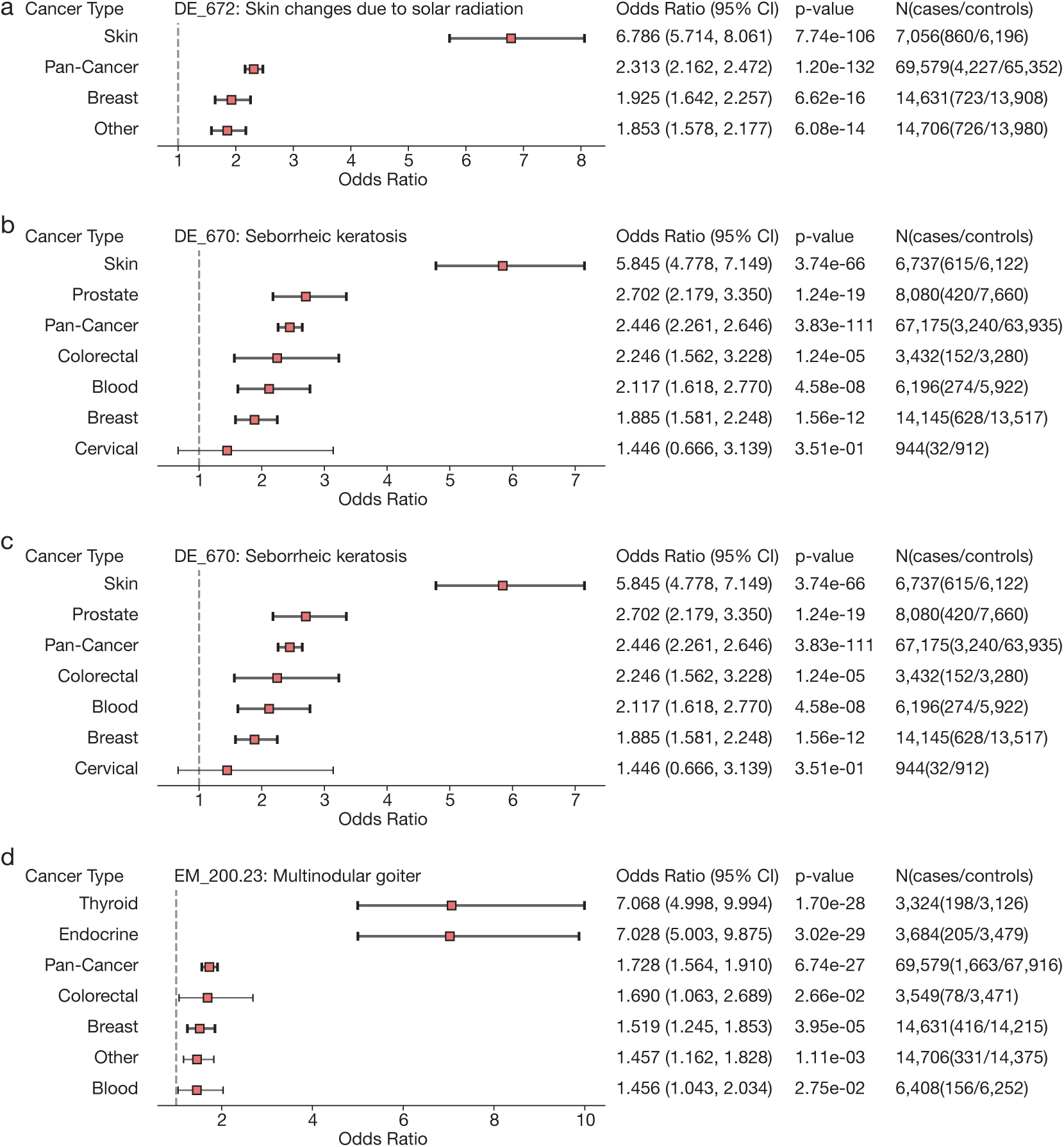
Cancer type specific risk associations for selected phenotypes with significant pan-cancer findings. Forest plots showing odds ratios (OR) and 95% confidence intervals (horizontal bars) for associations between individual phenotypes and specific cancer types. Each panel displays one phenotype across cancer types (rows), with pan-cancer estimate shown for reference. Vertical dashed line indicates OR = 1.0 (no association); values to the right indicate increased risk, values to the left indicate inverse associations. Bold text indicates Bonferroni-corrected significance. Case and control counts displayed in right column. **a**, Skin changes due to solar radiation (DE_672). **b**, Seborrheic keratosis (DE_670). **c**, Transplanted organ (SS_847). **d**, Multinodular goiter (EM_200.23).

**Fig 5.**
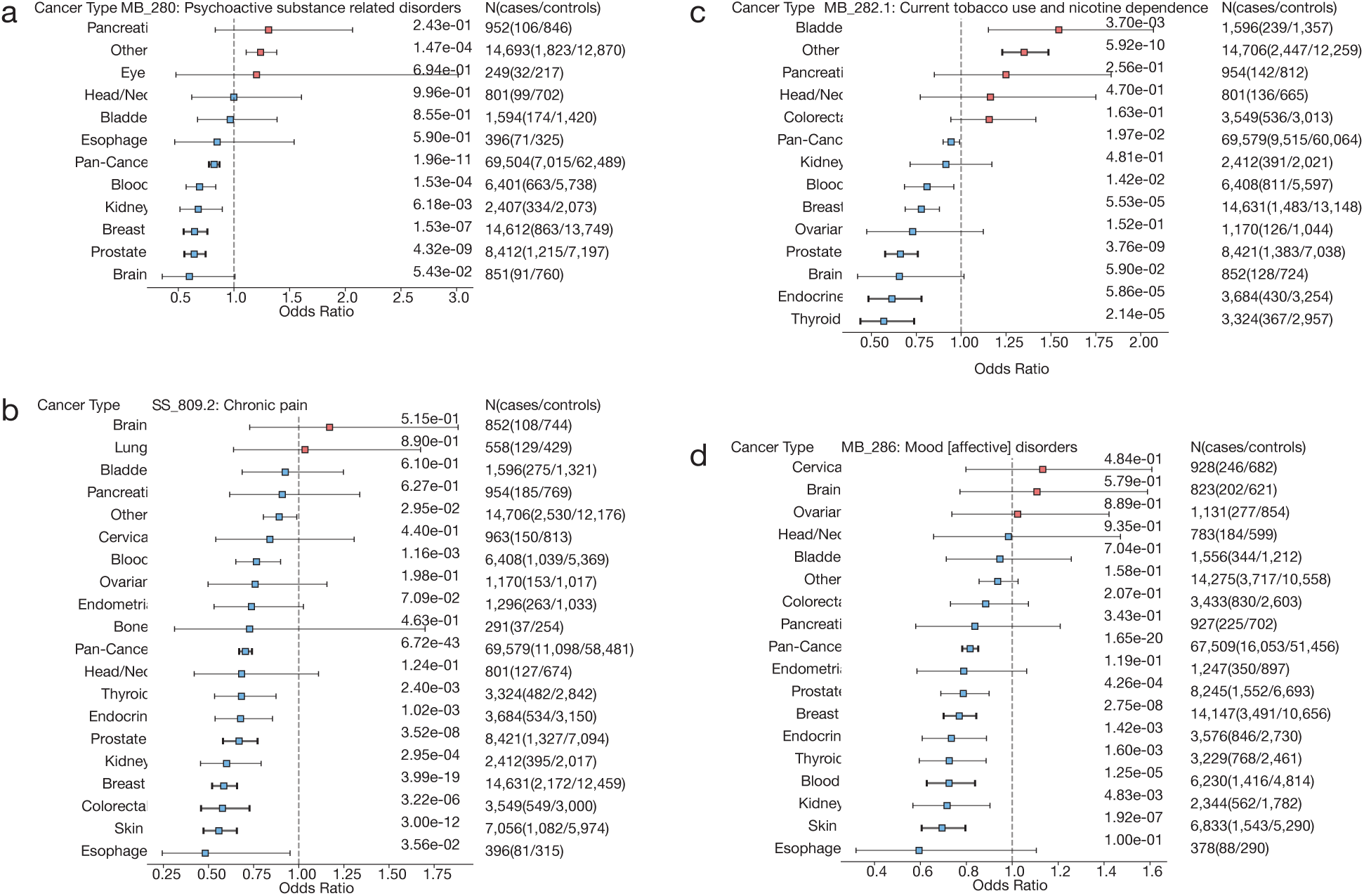
Cancer type specific inverse associations for selected phenotypes with significant pan-cancer findings. Forest plots showing odds ratios (OR) and 95% confidence intervals (horizontal bars) for associations between individual phenotypes and specific cancer types. Each panel displays one phenotype across cancer types (rows), with pan-cancer estimate shown for reference. Vertical dashed line indicates OR = 1.0 (no association); values to the right indicate increased risk, values to the left indicate inverse associations. Bold text indicates Bonferroni-corrected significance. Case and control counts displayed in right column. **a**, Psychoactive substance related disorders (MB_280). **b**, Chronic pain (SS_809.2). **c**, Current tobacco use and nicotine dependence (MB_282.1). **d**, Mood [affective] disorders (MB_286).

## Discussion

### Key Findings

In this study, we identified 232 statistically significant associations between clinical phenotypes at least 1 year before cancer diagnosis in a diverse cohort of over 69,000 individuals from *All of Us*. A central challenge in cancer epidemiology is the inherent heterogeneity of the disease. While cancer is frequently discussed as a singular entity, it encompasses a vast collection of distinct pathological conditions, each defined by unique evolutionary trajectories and etiologies.^22^ Our decision to perform PheWAS across 21 cancer types, rather than relying solely on pan-cancer analysis, enables identification of cancer type specific associations that may be obscured in aggregated analyses.

Established risk markers were confirmed across multiple cancer types. In females, breast conditions including benign mammary dysplasias (OR = 1.60, 95% CI: 1.48-1.73) and lump or mass in breast or nonspecific abnormal breast exam (OR = 1.45, 95% CI: 1.38-1.53) showed positive associations with subsequent cancer diagnosis, consistent with meta-analyses demonstrating that benign breast disease may confer elevated cancer risk.^23,24^ Thyroid disorders, including multinodular goiter (OR = 1.73, 95% CI: 1.56-1.91), were associated with cancer diagnosis, potentially reflecting shared endocrine dysregulation across multiple cancer types.^25–27^ In males, elevated prostate-specific antigen (OR = 2.92, 95% CI: 2.65-3.23) and benign prostatic hyperplasia (OR = 1.42, 95% CI: 1.33-1.53) confirmed well-documented markers of prostate cancer risk.^28,29^ Blood cancer analyses revealed strong associations with immune-related phenotypes, including monoclonal gammopathy (OR = 11.19, 95% CI: 6.31-19.85), reflecting the direct involvement of immune cells in hematologic malignancies. Kidney cancer showed a notable association with type 2 diabetes (OR = 1.71, 95% CI: 1.39-2.11), consistent with recent meta-analyses concluding that diabetes is an independent risk factor for the development of kidney cancer.^30^

Race stratified analyses revealed both shared risk factors and population-specific disease associations. Among Non-Hispanic Black participants, multiple anemia phenotypes emerged as significant pre-diagnosis correlates of cancer, including anemia and deficiency anemias. Prior research has established anemia as a risk factor for various malignancies, with recent evidence suggesting ethnic differences in anemia-cancer associations.^31,32^ For Hispanic participants, increased risk relating to Nonalcoholic Fatty Liver Disease (NAFLD) align with well-documented health disparities showing that Hispanic populations experience disproportionately high rates of NAFLD and may be partially explained by genetic susceptibility.^33,34^ The PNPLA3 I148M variant (rs738409), which impairs hepatic lipid metabolism and drives NAFLD progression, is highly prevalent in Hispanic populations.^35^ For Non-Hispanic White participants, dermatological findings such as actinic keratosis (OR = 2.62, 95% CI: 2.42-2.83) are consistent with the higher incidence of skin malignancies in North American and European populations.^36^

Inverse associations between phenotypes such as chronic pain and psychiatric conditions appear intriguing but require careful interpretation. Upon examination of baseline proportions, cancer cases exhibited a higher prevalence of both chronic pain (16.2% vs. 15.8%; unadjusted OR = 1.03) and mood disorders (24.1% vs. 22.5%; unadjusted OR = 1.09) compared to controls. However, after adjusting for condition count in our regression models, cancer was paradoxically associated with lower odds of both chronic pain (OR = 0.70, 95% CI: 0.67-0.74) and mood disorders (OR = 0.82, 95% CI: 0.78-0.85) across all four quartiles of condition count stratification. This pattern raises concerns about potential collider adjustment bias. When two variables independently influence a third variable, and that third variable is adjusted for, this can induce collider bias, which distorts observed associations.^37,38^ However, it is critical to distinguish between collider selection bias (inherent to who appears in EHR data) and collider adjustment bias (induced by including variables in regression models). While the former arises from restricting analyses to individuals with EHR records, the latter occurs when adjusting for a variable that is causally downstream of both exposure and outcome.^39^

In our study, it is essential to adjust for condition count to account for differential healthcare utilization between cases and controls.^40^ However, it is possible that both cancer and conditions such as chronic pain or mood disorders independently increase healthcare engagement and influence overall condition count. If so, then adjusting for it in regression models may induce spurious protective associations, similar to Simpson’s paradox where associations reverse direction across strata.^41^ This phenomenon parallels findings from Richardson et al., who reanalyzed UK Biobank data on neuroticism and mortality.^42^ The original study observed that neuroticism was associated with higher mortality in unadjusted analyses, but after adjusting for self-rated health, neuroticism appeared protective. Richardson et al. demonstrated that this reversal likely reflected collider bias: both neuroticism and mortality risk factors independently influence self-rated health, and conditioning on it induced spurious protective associations. Similarly, our observed protective associations for chronic pain and mood disorders emerged after adjusting for condition count, suggesting a comparable mechanism.

Importantly, collider bias is often small in magnitude, and controlling for confounding effects generally takes precedence over avoiding collider bias.^43–45^ We do not propose abandoning adjustment for condition count, as failing to control for healthcare utilization would introduce severe confounding.^45^ Rather, we propose that certain phenotype classes strongly associated with healthcare-seeking behavior may be especially susceptible to collider adjustment bias when condition count is included in models. The consistent pattern of protective associations across most psychiatric conditions suggests that interpretation should proceed with caution.

### Future Directions

These findings underscore the complexity of EHR-based susceptibility analyses. Future work should explore sensitivity analyses that assess the robustness of protective associations to different adjustment strategies, examine temporality of diagnosis patterns, and consider alternative methods for controlling healthcare utilization. Further investigation into how collider adjustment bias manifests in EHR-based studies represents an important direction for methodological research in precision medicine.

While *All of Us* provides unprecedented diversity and sample size, participants with extensive EHR data represent healthcare-engaged populations where conditions that drive medical utilization are enriched. Future population-based prospective cohort studies are essential for validating biobank findings and establishing relationships between clinical phenotypes and cancer risk. Beyond validation in prospective cohorts, replication of these findings in independent biobank resources is necessary to assess generalizability across healthcare systems and populations. Recent advances in machine learning models trained on EHR data demonstrate the potential for using longitudinal clinical trajectories for predicting disease onset, progression, and treatment response.^46^ However, such models trained on single healthcare system data, even when spanning millions of patients, face inherent limitations in generalizability. Predictive models trained on healthcare-engaged populations may fail to capture variations in practice patterns, population demographics, and access across different systems. The promise of computational medicine using EHR data is to be tempered by recognition that predictions reflect the specific patient populations and documentation practices of training institutions.

## Limitations

This study has several limitations inherent to EHR-based research. Diagnosis codes may be subject to inaccuracies related to billing practices, and true diagnoses may go unreported or unrecorded by physicians, potentially leading to phenotype misclassification.^47^ While we implemented matching and adjusted for key covariates, our retrospective cohort design using existing EHR data may introduce selection biases and residual mediating relationships remain to be fully understood.^48^ The PheWAS framework tests each phenotype independently, which, while enabling agnostic discovery, does not capture potential interactions between phenotypes. More sophisticated modeling may be necessary to reveal the multivariate complexity of disease development. Despite these limitations, disease-disease PheWAS remains valuable for hypothesis generation when methodological considerations are appropriately addressed.

## Conclusions

This study leveraged the diversity of *All of Us* to examine how clinical phenotypes prior to cancer diagnosis vary across cancer types and racial groups. Using EHR data from over 69,000 participants, we identified 232 pre-diagnosis associations in pan-cancer analyses, with substantial heterogeneity across 21 cancer-specific and race stratified subgroups. Our findings validate known cancer risk factors at scale, establishing a framework for susceptibility research in diverse biobank populations and demonstrating the power of inclusive datasets for precision medicine.

## Methods

### Overview: Study Population and Data Source

This study used data from the *All of Us* Controlled Tier Dataset version 8 (data cutoff: October 1, 2023; released: February 4, 2025).^14^ We included *All of Us* participants with EHR data, documented sex at birth (female or male), and self-reported race information from surveys.^49^ Analysis was restricted to unrelated individuals, cancer cases with EHR data at least 1 year pre-diagnosis, and controls without any malignant-related diagnoses to ensure clear case-control distinction (*n* = 235,430). To address age-related confounders inherent in longitudinal EHR data, we implemented a two-stage matched case-control design, resulting in a final cohort of 23,193 cases and 46,386 controls for PheWAS analysis (Fig. 1).^50,51^ For coding aspects of this study, Claude by Anthropic was used for debugging and troubleshooting.^52^

### Phenotyping

To identify individuals with a malignant neoplasm diagnosis, we used a SNOMED CT codelist adapted from Aschebrook-Kilfoy et al., who established a comprehensive cancer classification framework for *All of Us*.^53^ Any individual with at least one SNOMED code from the *All of Us* condition_occurrence table was considered a cancer case.^54^ Within *All of Us* version 8, we identified 55,800 unique cancer cases based on 24 SNOMED CT codes. Because our PheWAS analyzes International Classification of Diseases (ICD) coded phenotypes, we restricted cancer cases to those whose SNOMED diagnosis had a corresponding ICD code mapped in the All of Us data model, ensuring cases and phenotypes were derived from the same coding vocabulary (*n* = 52,676). After restricting to those with compatible ICD data in their EHR, 43,716 SNOMED-identified cancer cases remained. We further required cases to have EHR data at least 1 year prior to cancer diagnosis to enable meaningful assessment of pre-diagnosis phenotypes (*n* = 29,806), summarized in Supplementary Table 1.

For PheWAS analysis, these were grouped into 21 cancer types as follows: bladder (93689003), blood [leukemia (93143009), non-Hodgkin’s lymphoma (118601006), myeloma (109989006)], bone (93725000), brain (93727008), breast (372137005), cervical [cervix (372024009)], colorectal [colon (93761005), rectum (93984006)], endocrine [endocrine gland (371983001)], endometrial [endometrium (93781006)], esophageal [esophagus (371984007)], eye (371986009), head and neck [oral cavity (372001002)], kidney (93849006), lung (93880001), ovarian [ovary (93934004)], pancreatic [pancreas (372003004)], prostate (93974005), skin (94047004), stomach (372014001), thyroid (94098005), and other [primary malignant neoplasm (372087000)].

### Matched Case-Control Design

Using the MatchIt package (version 4.7.2) in R, cancer cases were first matched to possible controls using exact matching on sex at birth and self-reported race, with distance-based matching on birth year to establish demographic comparability and create a temporal index date for EHR censoring.^55^

We used nearest neighbor matching without replacement and variable ratio matching (target ratio of 4:1 controls per case). To prevent systematic bias in the matching order, we specified random matching order (m.order = “random”).^56^ Propensity scores for distance-based matching were estimated via logistic regression incorporating birth year. We imposed calipers of ±5 years on birth year to maximize sample retention. The matching procedure targeted the average treatment effect on the treated (ATT), enabling inferences specifically about the cancer patient population.^57^

Demographic balance was assessed using standardized mean differences (SMDs), variance ratios, and empirical cumulative distribution function (eCDF) statistics, with sufficient balance defined as SMD <0.1 for all demographics.^57^ Before matching, substantial imbalance existed between cases and controls, particularly for birth year (SMD = −1.14) and self-reported race categories (SMDs ranging from −0.17 for Non-Hispanic Black to 0.39 for Non-Hispanic White). After matching, balance improved markedly across demographics: birth year (SMD = −0.02), with perfect balance achieved for sex at birth and self-reported race categories (SMD = 0.0) due to exact matching. The maximum eCDF difference decreased from 0.38 to 0.02 for the distance metric post-matching, demonstrating improved distributional similarity.

Matching successfully paired cancer cases (*n* = 29,803) to controls (*n* = 108,036) with an average ratio of 3.63:1 due to the constraints of exact matching on sex at birth and self-reported race combined with the ±5 year caliper. Cases without matches (*n* = 3) represented individuals with extreme birth years or rare categories for whom suitable controls within caliper constraints were unavailable.

### EHR Censoring

For cancer cases, only pre-diagnosis EHR data is analytically relevant for identifying pre-existing conditions associated with subsequent cancer development. However, controls without cancer diagnoses have their entire EHR available for analysis, often extending years or decades beyond the timepoint when their matched case was diagnosed. This asymmetry creates spurious associations: controls can accumulate age-related diagnoses and era-specific conditions (e.g., COVID-19 diagnoses after 2020) that cases diagnosed earlier could never have accrued in their pre-diagnosis records. An example sensitivity analysis without control censoring is presented in Extended Data Fig. 1.

To address this bias, we implemented a matched censoring strategy ensuring that cases and controls are at risk for outcomes during the same age ranges. Controls were assigned an index date corresponding to their matched case’s diagnosis date. For both cases and controls, only EHR records occurring at least 1 year prior to the index date were retained for analysis, allowing assessment of pre-existing conditions rather than cancer-related symptoms. Following EHR censoring, controls that lacked any remaining pre-diagnosis EHR data (*n* = 40,935) were excluded from subsequent analysis. The resulting cohort (*n* = 96,904) consisted exclusively of cases (*n* = 29,803) and controls (*n* = 67,101) with pre-diagnosis EHR data.

### Post-Censoring Control Selection and Demographic Balance

Variable ratio matching (1:4 target) resulted in cases matched to between 1 and 5 controls in the post-censoring cohort. However, the overall imbalance in birth year distribution between cases and controls in the eligible population (case mean: 1955.7; control mean: 1970.5) created systematic differences in matching success. Cases who successfully matched to 4-5 controls represented a younger population, whereas older cases typically matched to only 1-2 controls due to the limited availability of similarly aged controls.

Consequently, birth year remained imbalanced in the post-censoring matched cohort (case mean: 1955.7; control mean: 1960.4), driven by the overrepresentation of younger, highly-matched cases (Fig. 1).

To address this residual imbalance while maintaining adequate statistical power, the post-censoring cohort (*n* = 96,904) was unmatched and returned to the matching pool. Second round matching employed 2:1 nearest neighbor matching without replacement. Matching specifications mirrored first round criteria with one addition: sex at birth (exact), self-reported race (exact), birth year (±5 year caliper), and age at diagnosis/index date (±5 year caliper). Propensity scores were estimated via logistic regression, with random matching order (m.order = “random”).

Before second round matching, residual imbalance persisted between cases and controls on birth year (SMD = −0.36). After second round matching, sufficient balance was achieved with all demographics obtaining a SMD <0.1: birth year (SMD = −0.09), age at diagnosis/censoring (SMD = −0.01); with perfect balance achieved for sex at birth and self-reported race categories (SMD = 0.0) due to exact matching, as presented in Extended Data Fig. 2, Extended Data Table 1.

Additional exclusion was applied to cases that had fewer than two controls remaining after second round matching (*n* = 6,610). The final analytic cohort (*n* = 69,579) comprised 23,193 cancer cases and 46,386 controls. Within the final cohort, demographic balance demonstrated substantial improvement across age-related metrics. Birth year achieved near-perfect balance (case mean: 1958.6; control mean: 1959.6), along with matched age at diagnosis/index date (case mean: 58.9 years; control mean: 58.9 years).

Median EHR observation windows (cases: 5.3 years; controls: 4.6 years), median unique diagnoses (cases: 28.0; controls 19.0), median total code occurrences (cases: 57.0; controls 36.0). Across the final cohort, we retained 82% of total ICD codes prior to cancer diagnosis (18,825,064/22,955,246) after filtering to 1 year before the matched age at diagnosis/index date (Fig. 1).

## Statistical Analysis

We implemented PheWAS to identify phenotypes associated with cancer using the PheTK package in Python.^40^ Clinical phenotypes were defined using the phecodeX phenotyping system, which maps ICD-9-CM and ICD-10-CM diagnosis codes from EHR data to standardized phenotype codes.^58^ ICD codes from the censored EHR records were mapped to phecodeX phenotypes using the PheTK Phecode module.^40^ Individuals were classified as cases for a given phecode if they had at least two occurrences on separate dates. Phecodes with fewer than 20 distinct cases were excluded from each analysis.PhecodeX organizes phenotypes into 18 major disease categories: Infections, Neoplasms, Blood/Immune, Endocrine/Metabolic, Mental, Neurological, Sense Organs, Cardiovascular, Respiratory, Gastrointestinal, Genitourinary, Dermatological, Musculoskeletal, Congenital, Symptoms, Neonatal, Pregnancy, and Genetic conditions.

Regression models were adjusted for potential confounders including sex at birth, self-reported race, age at diagnosis/index date, and two censored EHR metrics: observation window and unique condition count. Odds ratios (OR), 95% confidence intervals, and p-values were estimated using logistic regression. For the primary pan-cancer analysis, 2,317 phecodes were tested after excluding phecodes with fewer than 20 cases. With 1,646 converged tests, Bonferroni correction was applied with a significance threshold of α = 0.05/1,646 = 3.04×10⁻⁵ (equivalent to -log₁₀(*p*-value) > 4.52).

To examine whether associations vary by demographic characteristics or cancer type, we conducted stratified analyses using the final matched cohort. Rather than re-matching for each sub-analysis, we maintained the matched case-control pairs and stratified the cohort by the variable of interest. Each cancer type, race, age, and sex stratified analysis applied its own Bonferroni correction based on the number of phecodes tested in that specific analysis. Only phecodes that achieved model convergence were retained.^59^

## Data and code availability

This study used data from the *All of Us* Research Program’s Controlled Tier Dataset v8, available to authorized users on the workbench. All analyses were done using Jupyter Notebooks on the *All of Us* workbench, with notebooks available upon request (https://www.researchallofus.org/data-tools/workbench/).

## Data Availability

This study used data from the All of Us Research Program Controlled Tier Dataset v8, available to authorized users on the workbench. All analyses were done using Jupyter Notebooks on the All of Us workbench, with notebooks available upon request.

https://workbench.researchallofus.org/

## Acknowledgments

The content is solely the responsibility of the authors and does not necessarily represent the official views of the National Institutes of Health. In addition, the All of Us Research Program would not be possible without the partnership of its wonderful participants.

Research reported in this publication was supported by the National Cancer Institute of the National Institutes of Health under Award Number R15CA293800.

## Author contributions

M.H.B. and M.F.D. supervised and guided data analysis. C.C.D.R. drafted and wrote the manuscript. M.H.B., B.A.B., and M.F.D. revised the manuscript. C.C.D.R. and M.H.B. generated the figures. E.J.B and A.B.B. provided analysis and scientific input. B.E.R., E.J.B, A.B.B., and J.L.M. contributed to manuscript editing.

## Declaration of generative AI

Claude by Anthropic was used to revise text for grammar, clarity, and length optimization. Importantly, all scientific content, hypotheses, analyses, and conclusions remain the original work of the authors.

## Supplementary Materials

Supplementary Materials include Supplementary Text, two Supplementary Figures, one Supplementary Table, and one Supplementary Data.

## Extended Data

**Extended Data Fig 1.**
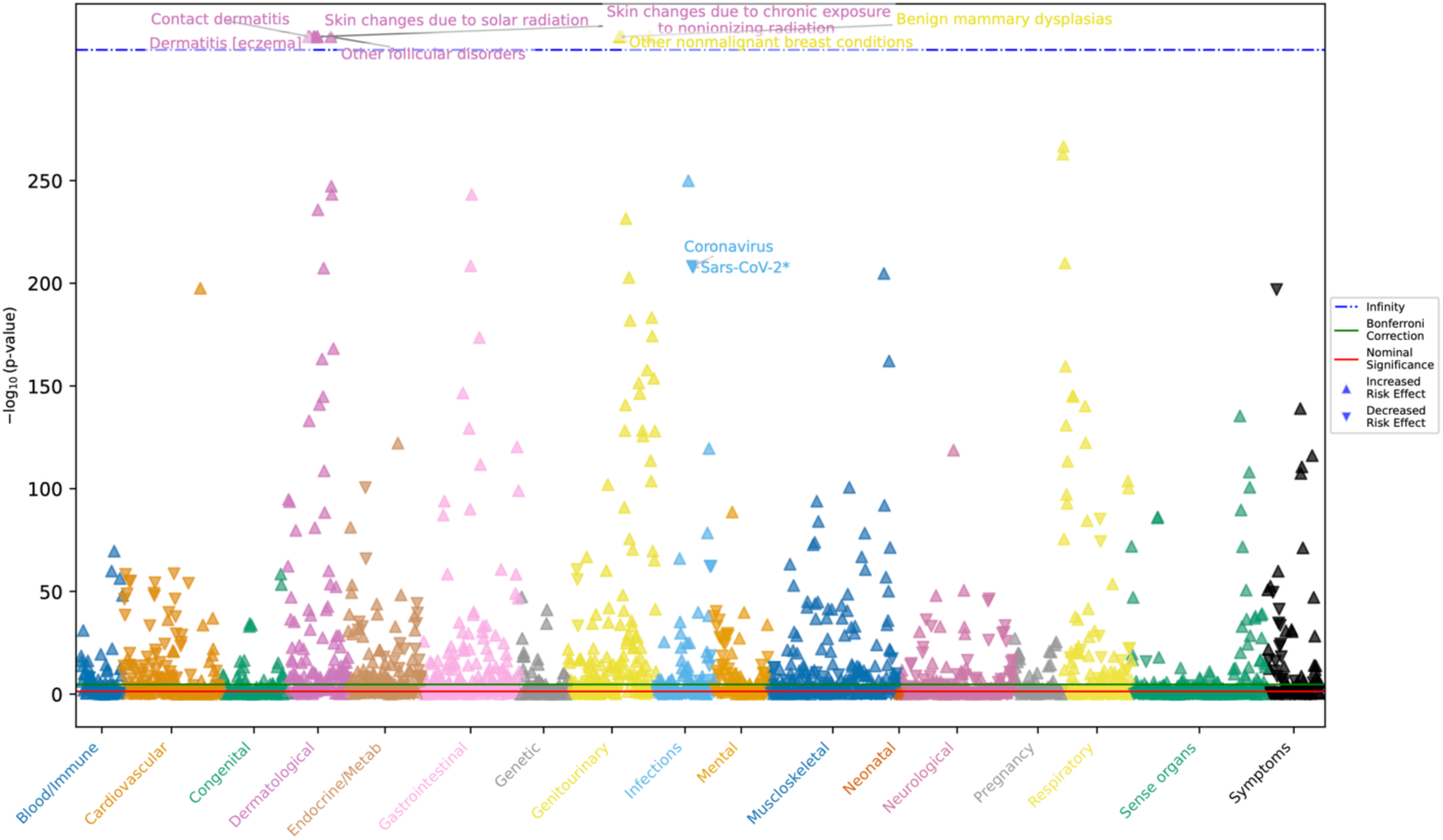
Age-related bias in uncensored EHR data. Manhattan plot showing −log₁₀(p-value) for associations between phecodeX phenotypes (x-axis, color-coded by disease category) and cancer diagnosis using the first-round matched cohort (cases: *n* = 29,803, controls: *n =* 108,036). Cases’ EHR data were restricted to pre-diagnosis records, while controls’ complete EHR histories were included, allowing controls to accumulate diagnoses years or decades beyond when their matched case was diagnosed. This uncensored analysis shows spurious associations driven by age-related confounding: controls exhibit enrichment for recent diagnoses (e.g., COVID-19 after 2020). These artifactual associations are addressed when proper temporal censoring is applied (see main analysis), demonstrating the necessity of matched observation windows for valid disease-disease association studies in longitudinal EHR data. Red horizontal line indicates p = 0.05; green line indicates Bonferroni-corrected significance threshold. Upward triangles represent increased risk associations; downward triangles represent inverse associations.

**Extended Data Fig 2.**
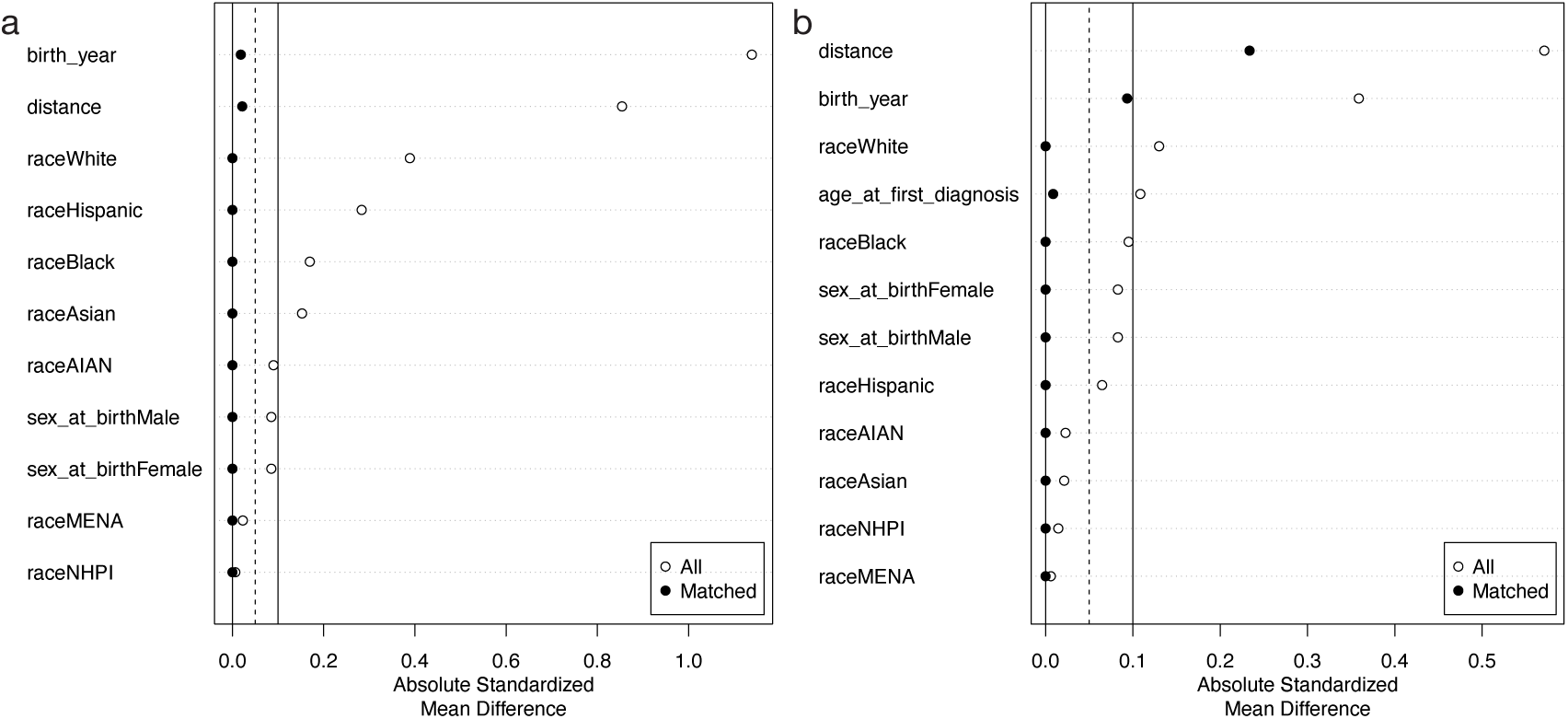
Demographic balance through two-stage matching. Absolute standardized mean differences (SMDs) before (open circles) and after (filled circles) matching for first-round and second-round matching procedures. **a**) shows balance improvement from initial unmatched cohort to first-round matched cohort using 4:1 nearest neighbor matching for birth year, with exact matching on sex at birth and self-reported race (cases: *n* = 29,803, controls: *n =* 108,036). **b**) demonstrates second-round matching performance on the post-censoring cohort. The final matched cohort (cases: *n* = 23,193, controls: *n =* 46,386) achieved SMDs <0.1 for all demographics. Vertical lines indicate SMD thresholds of 0.1 (solid) and 0.05 (dashed). Sex at birth and self-reported race achieved perfect balance (SMD = 0.00) through exact matching in both rounds.

**Extended Data Table 1.** Demographic Balance Across Matching Stages. Standardized mean differences (SMD), variance ratios, and empirical cumulative distribution function (eCDF) maximum differences for all demographics across four stages: unmatched cohort, first-round matching, post-censoring cohort, and second-round matching. SMD <0.1 indicates sufficient balance. Variance ratios between 0.5-2.0 indicate adequate distributional similarity. eCDF Max <0.05 indicates excellent distributional balance. Em dashes indicate variable not applicable at that stage (age at diagnosis/index date only exists after EHR censoring). Sex at birth and self-reported race achieved perfect balance (SMD = 0.00) through exact matching in both rounds. AIAN = American Indian/Alaska Native; MENA = Middle Eastern/North African; NHPI = Native Hawaiian/Pacific Islander.

| Demographic | Unmatched |  |  | 1st Round Match |  |  | Post-Censoring |  |  | 2nd Round Match |  |  |
| --- | --- | --- | --- | --- | --- | --- | --- | --- | --- | --- | --- | --- |
|  | SMD | Var Ratio | eCDF Max | SMD | Var Ratio | eCDF Max | SMD | Var Ratio | eCDF Max | SMD | Var Ratio | eCDF Max |
| Propensity Score Distance | 0.85 | 1.33 | 0.38 | 0.02 | 1.01 | 0.02 | 0.57 | 2.41 | 0.26 | 0.23 | 1.51 | 0.12 |
| Birth Year | -1.14 | 0.62 | 0.38 | -0.02 | 1.01 | 0.02 | -0.36 | 1.11 | 0.19 | -0.09 | 1.09 | 0.07 |
| Age at Diagnosis/<br>Index Date | — | — | — | — | — | — | 0.11 | 1.13 | 0.07 | -0.01 | 1.05 | 0.01 |
| Female | -0.09 | — | 0.04 | 0.00 | — | 0.00 | -0.08 | — | 0.04 | 0.00 | — | 0.00 |
| Male | 0.09 | — | 0.04 | 0.00 | — | 0.00 | 0.08 | — | 0.04 | 0.00 | — | 0.00 |
| Non-Hispanic White | 0.39 | — | 0.17 | 0.00 | — | 0.00 | 0.13 | — | 0.06 | 0.00 | — | 0.00 |
| Non-Hispanic Black | -0.17 | — | 0.06 | 0.00 | — | 0.00 | -0.10 | — | 0.03 | 0.00 | — | 0.00 |
| Hispanic | -0.28 | — | 0.09 | 0.00 | — | 0.00 | -0.06 | — | 0.02 | 0.00 | — | 0.00 |
| Asian | -0.15 | — | 0.02 | 0.00 | — | 0.00 | -0.02 | — | 0.00 | 0.00 | — | 0.00 |
| AIAN | -0.09 | — | 0.01 | 0.00 | — | 0.00 | -0.02 | — | 0.00 | 0.00 | — | 0.00 |
| MENA | -0.02 | — | 0.00 | 0.00 | — | 0.00 | -0.01 | — | 0.00 | 0.00 | — | 0.00 |
| NHPI | -0.01 | — | 0.00 | 0.00 | — | 0.00 | -0.01 | — | 0.00 | 0.00 | — | 0.00 |

## References

1. Collins FS, Varmus H. A New Initiative on Precision Medicine. N Engl J Med. 2015;372(9):793–795. doi:10.1056/NEJMp1500523

2. Ashley EA. Towards precision medicine. Nat Rev Genet. 2016;17(9):507–522. doi:10.1038/nrg.2016.86

3. The “All of Us” Research Program. N Engl J Med. 2019;381(7):668–676. doi:10.1056/NEJMsr1809937

4. Bycroft C, Freeman C, Petkova D, et al. The UK Biobank resource with deep phenotyping and genomic data. Nature. 2018;562(7726):203–209. doi:10.1038/s41586-018-0579-z

5. Abdellaoui A, Sanchez-Roige S, Sealock J, et al. Phenome-wide investigation of health outcomes associated with genetic predisposition to loneliness. Hum Mol Genet. 2019;28(22):3853–3865. doi:10.1093/hmg/ddz219

6. Li S, Schooling CM. A phenome-wide association study of ABO blood groups. BMC Med. 2020;18:334. doi:10.1186/s12916-020-01795-4

7. Cancer Facts & Figures 2026. Published online 1930.

8. Arem H, Loftfield E. Cancer Epidemiology: A Survey of Modifiable Risk Factors for Prevention and Survivorship. Am J Lifestyle Med. 2017;12(3):200–210. doi:10.1177/1559827617700600

9. Marino P, Mininni M, Deiana G, et al. Healthy Lifestyle and Cancer Risk: Modifiable Risk Factors to Prevent Cancer. Nutrients. 2024;16(6):800. doi:10.3390/nu16060800

10. Reducing the Risk of Cancer Development - CPR25. Cancer Progress Report. Accessed February 11, 2026. https://cancerprogressreport.aacr.org/progress/cpr25-contents/cpr25-reducing-the-risk-of-cancer-development/

11. Spratt DE, Chan T, Waldron L, et al. Racial/Ethnic Disparities in Genomic Sequencing. JAMA Oncol. 2016;2(8):1070–1074. doi:10.1001/jamaoncol.2016.1854

12. Nugent A, Conatser KR, Turner LL, Nugent JT, Sarino EMB, Ricks-Santi LJ. Reporting of race in genome and exome sequencing studies of cancer: a scoping review of the literature. Genet Med. 2019;21(12):2676–2680. doi:10.1038/s41436-019-0558-2

13. Yuan J, Hu Z, Mahal BA, et al. Integrated Analysis of Genetic Ancestry and Genomic Alterations across Cancers. Cancer Cell. 2018;34(4):549–560.e9. doi:10.1016/j.ccell.2018.08.019

14. Curated Data Repository (CDR) version 8 Release Notes. User Support. February 4, 2025. Accessed February 11, 2026. https://support.researchallofus.org/hc/en-us/articles/30294451486356-Curated-Data-Repository-CDR-version-8-Release-Notes

15. Weinstein JN, Collisson EA, Mills GB, et al. The Cancer Genome Atlas Pan-Cancer Analysis Project. Nat Genet. 2013;45(10):1113–1120. doi:10.1038/ng.2764

16. Tomasetti C, Vogelstein B. Variation in cancer risk among tissues can be explained by the number of stem cell divisions. Science. 2015;347(6217):78–81. doi:10.1126/science.1260825

17. Pendergrass SA, Brown-Gentry K, Dudek SM, et al. The Use of Phenome-Wide Association Studies (PheWAS) for Exploration of Novel Genotype-Phenotype Relationships and Pleiotropy Discovery. Genet Epidemiol. 2011;35(5):410–422. doi:10.1002/gepi.20589

18. Diogo D, Tian C, Franklin CS, et al. Phenome-wide association studies across large population cohorts support drug target validation. Nat Commun. 2018;9:4285. doi:10.1038/s41467-018-06540-3

19. Salvatore M, Gu T, Mack JA, et al. A Phenome-Wide Association Study (PheWAS) of COVID-19 Outcomes by Race Using the Electronic Health Records Data in Michigan Medicine. J Clin Med. 2021;10(7):1351. doi:10.3390/jcm10071351

20. Kerley CI, Chaganti S, Nguyen TQ, et al. pyPheWAS: A Phenome-Disease Association Tool for Electronic Medical Record Analysis. Neuroinformatics. 2022;20(2):483–505. doi:10.1007/s12021-021-09553-4

21. Chaganti S, Mawn LA, Kang H, et al. Electronic Medical Record Context Signatures Improve Diagnostic Classification using Medical Image Computing. IEEE J Biomed Health Inform. 2019;23(5):2052–2062. doi:10.1109/JBHI.2018.2890084

22. Hoadley KA, Yau C, Hinoue T, et al. Cell-of-Origin Patterns Dominate the Molecular Classification of 10,000 Tumors from 33 Types of Cancer. Cell. 2018;173(2):291–304.e6. doi:10.1016/j.cell.2018.03.022

23. Dyrstad SW, Yan Y, Fowler AM, Colditz GA. Breast cancer risk associated with benign breast disease: systematic review and meta-analysis. Breast Cancer Res Treat. 2015;149(3):569–575. doi:10.1007/s10549-014-3254-6

24. Luo J, Hendryx M, Nassir R, Cheng TYD, Lane D, Margolis KL. Benign breast disease and risk of thyroid cancer. Cancer Causes Control. 2017;28(9):913–920. doi:10.1007/s10552-017-0918-7

25. Belfiore A, La Rosa GL, La Porta GA, et al. Cancer risk in patients with cold thyroid nodules: relevance of iodine intake, sex, age, and multinodularity. Am J Med. 1992;93(4):363–369. doi:10.1016/0002-9343(92)90164-7

26. Hegedüs L. The Thyroid Nodule. New England Journal of Medicine. 2004;351(17):1764–1771. doi:10.1056/NEJMcp031436

27. Yildirim Simsir I, Cetinkalp S, Kabalak T. Review of Factors Contributing to Nodular Goiter and Thyroid Carcinoma. Med Princ Pract. 2020;29(1):1–5. doi:10.1159/000503575

28. Guo J, Zhou L, He J, et al. Benign prostatic hyperplasia and risk of urological cancers: a prospective cohort study based on the UK biobank. Discov Oncol. 2025;16:1771. doi:10.1007/s12672-025-03564-2

29. Carter HB, Metter EJ, Wright J, Landis P, Platz E, Walsh PC. Prostate-specific antigen and all-cause mortality: results from the Baltimore Longitudinal Study On Aging. J Natl Cancer Inst. 2004;96(7):557–558. doi:10.1093/jnci/djh111

30. Bonilla-Sanchez A, Rojas-Munoz J, Garcia-Perdomo HA. Association Between Diabetes and the Risk of Kidney Cancer: Systematic Review and Meta-Analysis. Clin Diabetes. 2022;40(3):270–282. doi:10.2337/cd21-0013

31. Hung N, Shen CC, Hu YW, et al. Risk of Cancer in Patients with Iron Deficiency Anemia: A Nationwide Population-Based Study. PLoS One. 2015;10(3):e0119647. doi:10.1371/journal.pone.0119647

32. Down L, Barlow M, Bailey SE, et al. Anaemia, ethnicity, and cancer incidence: a retrospective cohort study in primary care. Br J Gen Pract. 75(759):e678–e685. doi:10.3399/BJGP.2024.0762

33. Carrion AF, Ghanta R, Carrasquillo O, Martin P. Chronic Liver Disease in the Hispanic Population of the United States. Clin Gastroenterol Hepatol. 2011;9(10):834–e110. doi:10.1016/j.cgh.2011.04.027

34. Samji NS, Snell PD, Singal AK, Satapathy SK. Racial Disparities in Diagnosis and Prognosis of Nonalcoholic Fatty Liver Disease. Clin Liver Dis (Hoboken). 2020;16(2):66–72. doi:10.1002/cld.948

35. Romeo S, Kozlitina J, Xing C, et al. Genetic variation in PNPLA3 confers susceptibility to nonalcoholic fatty liver disease. Nat Genet. 2008;40(12):1461–1465. doi:10.1038/ng.257

36. Karimkhani C, Green AC, Nijsten T, et al. The global burden of melanoma: results from the Global Burden of Disease Study 2015. Br J Dermatol. 2017;177(1):134–140. doi:10.1111/bjd.15510

37. Holmberg MJ, Andersen LW. Collider Bias. JAMA. 2022;327(13):1282–1283. doi:10.1001/jama.2022.1820

38. Example of collider bias. Accessed May 19, 2026. https://jeremydfoote.com/notebooks/collider.html

39. Lu H, Gonsalves GS, Westreich D. Selection Bias Requires Selection: The Case of Collider Stratification Bias. Am J Epidemiol. 2023;193(3):407–409. doi:10.1093/aje/kwad213

40. Tran TC, Schlueter DJ, Zeng C, Mo H, Carroll RJ, Denny JC. PheWAS analysis on large-scale biobank data with PheTK. Bioinformatics. 2024;41(1):btae719. doi:10.1093/bioinformatics/btae719

41. Simpson EH. The Interpretation of Interaction in Contingency Tables. Royal Statistical Society Journal Series B: Methodological. 1951;13(2):238–241. doi:10.1111/j.2517-6161.1951.tb00088.x

42. Richardson TG, Davey Smith G, Munafò MR. Conditioning on a Collider May Induce Spurious Associations: Do the Results of Gale et al. (2017) Support a Health-Protective Effect of Neuroticism in Population Subgroups? Psychol Sci. 2019;30(4):629–632. doi:10.1177/0956797618774532

43. Liu W, Brookhart MA, Schneeweiss S, Mi X, Setoguchi S. Implications of M bias in epidemiologic studies: a simulation study. Am J Epidemiol. 2012;176(10):938–948. doi:10.1093/aje/kws165

44. Ding P, Miratrix LW. To Adjust or Not to Adjust? Sensitivity Analysis of M-Bias and Butterfly-Bias. Journal of Causal Inference. 2015;3(1):41–57. doi:10.1515/jci-2013-0021

45. Bhatt N. Covariate Selection in Causal Inference: Good and Bad Controls. Medium. July 29, 2025. Accessed May 19, 2026. https://booking.ai/covariate-selection-in-causal-inference-good-and-bad-controls-5f56126a984a

46. Zhang A, Ding T, Wagner SJ, et al. A multimodal and temporal foundation model for virtual patient representations at healthcare system scale. arXiv. Preprint posted online April 21, 2026:arXiv:2604.18570. doi:10.48550/arXiv.2604.18570

47. Casey JA, Schwartz BS, Stewart WF, Adler NE. Using Electronic Health Records for Population Health Research: A Review of Methods and Applications. Annu Rev Public Health. 2016;37:61–81. doi:10.1146/annurev-publhealth-032315-021353

48. Weiskopf NG, Dorr DA, Jackson C, Lehmann HP, Thompson CA. Healthcare utilization is a collider: an introduction to collider bias in EHR data reuse. J Am Med Inform Assoc. 2023;30(5):971–977. doi:10.1093/jamia/ocad013

49. Ramirez AH, Sulieman L, Schlueter DJ, et al. The All of Us Research Program: Data quality, utility, and diversity. Patterns (N Y). 2022;3(8):100570. doi:10.1016/j.patter.2022.100570

50. Yuan W, Beaulieu-Jones BK, Yu KH, et al. Temporal bias in case-control design: preventing reliable predictions of the future. Nat Commun. 2021;12:1107. doi:10.1038/s41467-021-21390-2

51. Lutsey PL. Case-control studies: Increasing scientific rigor in control selection. Res Pract Thromb Haemost. 2023;7(2):100090. doi:10.1016/j.rpth.2023.100090

52. Claude 4.5 Sonnet [Large language model]. https://claude.ai/

53. Aschebrook-Kilfoy B, Zakin P, Craver A, et al. An Overview of Cancer in the First 315,000 All of Us Participants. PLOS ONE. 2022;17(9):e0272522. doi:10.1371/journal.pone.0272522

54. Bates BA, Bates KE, Boris SA, et al. Intersection of rare pathogenic variants from TCGA in the All of Us Research Program v6. HGGADVANCE. 2025;6(2). doi:10.1016/j.xhgg.2025.100405

55. Ho D, Imai K, King G, Stuart E, Whitworth A, Greifer N. MatchIt: Nonparametric Preprocessing for Parametric Causal Inference. Published online May 30, 2025. Accessed February 11, 2026. https://cran.r-project.org/web/packages/MatchIt/index.html

56. Austin PC. A comparison of 12 algorithms for matching on the propensity score. Statistics in Medicine. 2014;33(6):1057–1069. doi:10.1002/sim.6004

57. Ho DE, Imai K, King G, Stuart EA. Matching as Nonparametric Preprocessing for Reducing Model Dependence in Parametric Causal Inference. Political Analysis. 2007;15(3):199–236. doi:10.1093/pan/mpl013

58. Shuey MM, Stead WW, Aka I, et al. Next-generation phenotyping: introducing phecodeX for enhanced discovery research in medical phenomics. Bioinformatics. 2023;39(11):btad655. doi:10.1093/bioinformatics/btad655

59. Clark RG, Blanchard W, Hui FKC, Tian R, Woods H. Dealing with complete separation and quasi-complete separation in logistic regression for linguistic data. Research Methods in Applied Linguistics. 2023;2(1):100044. doi:10.1016/j.rmal.2023.100044

