## Supplementary Material for "Phenome-Wide Association Study of Pre-Cancer Diagnosis Electronic Health Records Identifies Risk and Inverse Associations in the All of Us Research Program"

Rich *et al.*

**This PDF file includes:**

Supplementary Text 1

Figs. S1 to S2

Supplementary Table 1

Supplementary Data 1

**Supplementary information**

**Supplementary Text 1 Detailed Matching Methodology.**

**Rationale for Nearest Neighbor Matching**

We selected nearest neighbor matching for its computational efficiency and comparable performance to more complex methods in most scenarios. Nearest neighbor matching offers straightforward interpretation and transparency in the matching process, while retaining flexibility to implement exact matching constraints (sex at birth, self-reported race) alongside distance-based matching (continuous variables). We implemented variable ratio matching with a target of 4:1 controls per case. This approach balances two competing objectives: maximizing statistical power through larger control pools while maintaining match quality. The MatchIt documentation notes diminishing returns in balance improvement beyond a 4:1 ratio, as additional matched controls contribute progressively less to variance reduction while potentially degrading average match quality.^1^ In our cohort, the actual achieved ratio varied (3.63:1 after first-round matching, 2:1 after second-round matching) due to constraints imposed by exact matching requirements and caliper restrictions.

**Matching Order Evaluation**

For nearest neighbor matching, the order in which units are matched can influence both the quality of individual matches and the final matched sample composition. We evaluated six matching order strategies to assess their impact on our cohort: largest (highest propensity score first), smallest (lowest propensity score first), closest (best pairwise matches first), farthest (worst pairwise matches first), random, and data order. Across these strategies, cohort sizes ranged from 128,817 to 138,878 individuals, with case counts varying from 25,041 to 30,844 and control counts from 104,776 to 109,034. The achieved case-control ratios ranged from 3.53:1 to 4,18:1. Importantly, no single ordering method produced dramatically superior results. Matching order effects, while present, did not fundamentally alter cohort composition in our dataset.

We selected random matching order (m.order = "random") for our final analysis to avoid introducing systematic bias through the matching sequence. Rubin (1973) and Austin (2013) both recommend random ordering as it prevents prioritization artifacts that could favor or disadvantage certain subgroups.^2,3^ For example, "largest" ordering would match cases with extreme propensity scores first, potentially creating selection bias if those cases differ from others on unmeasured characteristics. Random ordering ensures that no subgroup receives preferential access to the best available matches.

**Caliper Selection and Justification**

Calipers define the maximum allowable distance between matched units on specified variables, preventing poor-quality matches that could introduce residual confounding. We imposed two calipers in our initial matching procedure: ±5 years on birth year and ±5 years on age at diagnosis/index date. The ±5 year caliper on birth year ensures cases and controls experienced similar historical contexts, healthcare systems, and age-related health trajectories. A 5-year window balances temporal similarity with practical matching feasibility; tighter windows would exclude too many cases, while wider windows would allow matches across meaningfully different cohort effects. For second-round matching, we added the ±5 years caliper on age at diagnosis/index date.

**Necessity and Implementation of Second-Round Matching**

After first-round matching and EHR censoring, substantial covariate imbalance persisted, necessitating a second matching round. The variable ratio matching strategy (target 4:1) created systematic age imbalance: younger cases successfully matched to 4–5 controls, while older cases matched to only 1–2 controls due to limited availability of similarly aged controls in the pool. This differential matching success overrepresented younger cases in the post-censoring cohort, with median birth year remaining imbalanced (cases: 1955 vs. controls: 1960).

Second-round matching directly addressed this issue by returning all cases and controls to the matching pool and implementing 2:1 nearest neighbor matching with enhanced specifications. This approach allowed us to preferentially retain cases with balanced matches while excluding cases whose matches were systematically poor. The final cohort (23,193 cases, 46,386 controls) demonstrated near-perfect balance (all SMDs <0.1) on all matching variables.

**Robustness and Methodological Implications**

Our iterative matching approach, combining temporal censoring with two-stage matched case-control design, addresses fundamental challenges in longitudinal EHR studies that previous PheWAS investigations have acknowledged but not fully resolved.^4^ Our methodology demonstrates that proper handling of temporal bias requires both temporal alignment (censoring) and population balance (matching), implemented in the correct sequence: initial demographic matching, temporal censoring, then refined matching on post-censoring characteristics. Future PheWAS studies using longitudinal EHR data should consider similar two-stage approaches, particularly when variable ratio matching is employed or when post-censoring imbalances emerge.

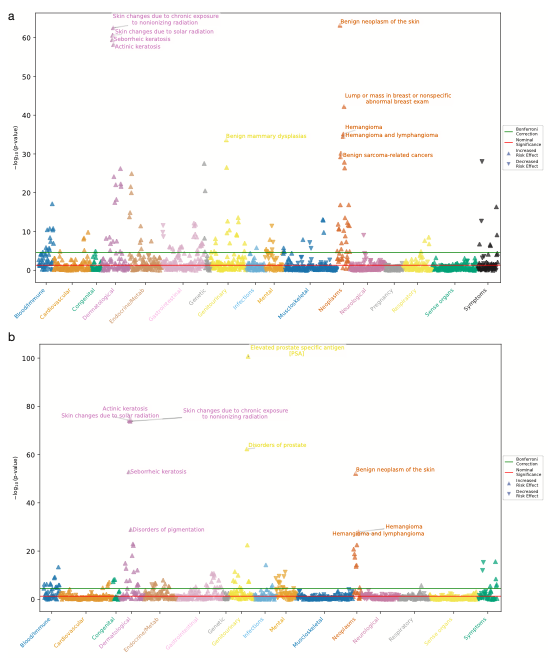

**Fig. S1. Sex Stratified PheWAS.**

Manhattan plots showing −log₁₀(p-value) for associations between phecodeX phenotypes (x-axis, color-coded by disease category) and cancer diagnosis. Red horizontal line indicates p = 0.05; green line indicates Bonferroni-corrected significance threshold. Upward triangles represent increased risk associations; downward triangles represent inverse associations. **a**) Female participants (cases: *n* = 14,607, controls: *n* = 29,214). **b**) Male participants (cases: *n* = 8,586, controls: *n* = 17,172). Each analysis applied independent Bonferroni correction based on phenotypes tested within that stratum.

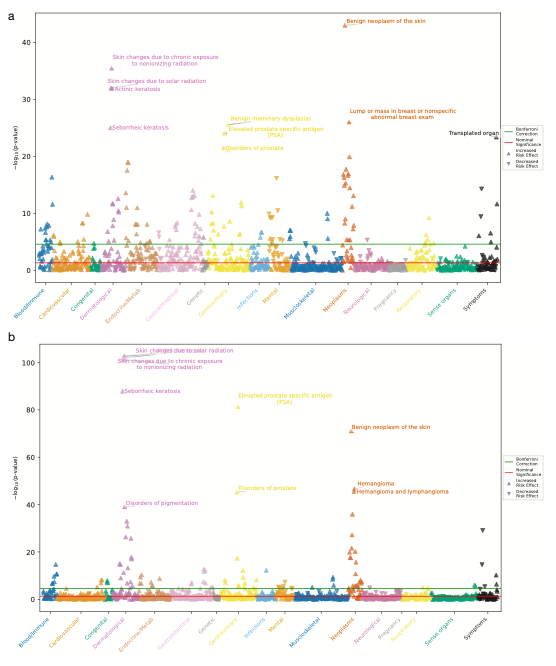

**Fig. S2. Age Stratified PheWAS.**

Manhattan plots showing −log₁₀(p-value) for associations between phecodeX phenotypes (x-axis, color-coded by disease category) and cancer diagnosis. Red horizontal line indicates p = 0.05; green line indicates Bonferroni-corrected significance threshold. Upward triangles represent increased risk associations; downward triangles represent inverse associations. **a**) Participants under 60 years old (cases: *n* = 11,0115, controls: *n* = 22,030). **b**) Participants over 60 years old (cases: *n* = 12,178, controls: *n* = 24,356). Each analysis applied independent Bonferroni correction based on phenotypes tested within that stratum.

| Cancer Type | SNOMED CT Code(s) | Specific Type | Original Cases | Final Cohort | Female | Male |
| --- | --- | --- | --- | --- | --- | --- |
| Bladder | 93689003 | bladder | 1,365 | 532 | 185 | 347 |
| Blood | 93143009, 118601006, 109989006 | leukemia, non-Hodgkin's lymphoma, myeloma | 4,964 | 2,136 | 1,220 | 916 |
| Bone | 93725000 | bone | 277 | 97 | 52 | 45 |
| Brain | 93727008 | brain | 790 | 284 | 182 | 102 |
| Breast | 372137005 | breast | 10,453 | 4,877 | 4,877 | 0 |
| Cervical | 372024009 | cervix | 787 | 321 | 321 | 0 |
| Colorectal | 93761005, 93984006 | colon, rectum | 2,745 | 1,183 | 704 | 479 |
| Endocrine | 371983001 | endocrine gland | 2,718 | 1,228 | 954 | 274 |
| Endometrial | 93781006 | endometrium | 743 | 432 | 432 | 0 |
| Esophageal | 371984007 | esophagus | 298 | 132 | 44 | 88 |
| Eye | 371986009 | eye | 170 | 83 | 46 | 37 |
| Head and Neck | 372001002 | oral cavity | 623 | 267 | 123 | 144 |
| Kidney | 93849006 | kidney | 1,696 | 804 | 367 | 437 |
| Lung | 93880001 | lung | 347 | 186 | 118 | 68 |
| Ovarian | 93934004 | ovary | 957 | 390 | 390 | 0 |
| Pancreatic | 372003004 | pancreas | 733 | 318 | 185 | 133 |
| Prostate | 93974005 | prostate | 7,045 | 2,809 | 0 | 2,809 |
| Skin | 94047004 | skin | 6,094 | 2,352 | 1,408 | 944 |
| Stomach | 372014001 | stomach | 285 | 122 | 73 | 49 |
| Thyroid | 94098005 | thyroid gland | 2,463 | 1,108 | 885 | 223 |
| Other | 372087000 | primary malignant neoplasm | 10,232 | 4,903 | 3,093 | 1,810 |

**Supplementary Table 1. Cancer Classification and Case Counts.**

Cancer Classification and Case Counts. Cancer cases were identified using SNOMED CT codes adapted from Aschebrook-Kilfoy et al. (2022). Original cases represent all individuals with at least one SNOMED cancer code in the All of Us version 8 dataset. Final cohort cases are those included in the PheWAS analysis after matching, EHR censoring, and quality filters. Note: Blood cancers combine leukemia, non-Hodgkin's lymphoma, and multiple myeloma; colorectal combines colon and rectum cancers. Individuals could have multiple cancer types; therefore, the sum of cancer type instances (55,785 original, 24,564 final) exceeds the number of unique persons (52,676 original, 23,193 final).

**Supplementary Data 1: Complete PheWAS Results.**

[Cancer_PheTK_Results.xlsx](https://docs.google.com/spreadsheets/d/1DxGerUhTjeHyB5229QeBPhO52-JP1JaC/edit?usp=sharing&ouid=116954012639217625924&rtpof=true&sd=true)

**References**

1.	Greifer N. MatchIt: Nonparametric Preprocessing for Parametric Causal Inference. R package version 4.7.2. 2025. https://cran.r-project.org/web/packages/MatchIt/vignettes/matching-methods.html

2.	Rubin DB. Matching to Remove Bias in Observational Studies. *Biometrics*. 1973;29(1):159-183. doi:10.2307/2529684

3.	Austin PC. The performance of different propensity score methods for estimating marginal hazard ratios. *Stat Med*. 2013;32(16):2837-2849. doi:10.1002/sim.5705

4.	Salvatore M, Gu T, Mack JA, et al. A Phenome-Wide Association Study (PheWAS) of COVID-19 Outcomes by Race Using the Electronic Health Records Data in Michigan Medicine. *J Clin Med*. 2021;10(7):1351. doi:10.3390/jcm10071351
